# Cold Hands, Warm Heart: Quality of Life, Wellbeing, and Mental Health in Raynaud’s Disease – An International Survey Study

**DOI:** 10.1101/2025.06.06.25329136

**Authors:** Eleftheria Vaportzis

## Abstract

**Objectives:** The study assessed the quality of life, wellbeing, and mental health of people living with Raynaud’s disease (RD). It employed an RD-specific measure for quality of life, used a large international sample, and included meteorological variables in the analyses. It also compared outcomes between primary and secondary RD and people without an RD diagnosis.

**Methods:** A cross-sectional design was used. Participants (n = 720) completed a demographics questionnaire, The Raynaud-Specific Quality of Life Questionnaire (RQLQ), WHO-5 Wellbeing Index and DASS-21. Meteorological variables were recorded based on the participants’ location.

**Results:** Participants with secondary RD reported lower quality of life and wellbeing, and higher anxiety, depression, and pain, than participants with primary RD or without a diagnosis. Participants without a diagnosis reported worse mental health than groups with a diagnosis. Participants in tropical climates reported the lowest quality of life, and those in temperate climates had the lowest wellbeing. Pain and symptom severity were the strongest predictors of quality of life (accounting for 30% of the variance), while meteorological variables accounted for up to 2% of the variance.

**Conclusion:** RD negatively affects quality of life, wellbeing, and mental health, particularly for people living with secondary RD. Pain and symptom severity are key determinants. Meteorological factors contribute minimally. Tailored interventions focusing on symptoms management should be prioritised. Meteorological influences should be considered in healthcare planning alongside clinical and psychosocial factors.

**Key messages:**

- Pain and symptom severity strongly predict quality of life in Raynaud’s disease
- Individuals without a diagnosis report worse mental health than individuals with a diagnosis
- Warmer climates do not necessarily protect against reduced qualify of life in Raynaud’s disease

## Introduction

Raynaud’s disease (RD) is a disorder of the peripheral vascular system characterised by episodes of pain, numbness, and colour changes in the extremities due to cold or stress. RD can be primary (pRD) or secondary (sRD) to an underlying condition such as scleroderma (1). The prevalence of pRD in the general population is around 5%, and although it varies by country (2), it carries a significant burden. It is estimated that pRD accounts for 80-90% of RD cases, and sRD accounts for 10-20% (3). Although some studies found the prevalence of RD to be higher in colder climates (4), other studies reported conflicting evidence (5, 6).

The impact of RD on quality of life depends on several factors, including the frequency, duration, and severity of episodes, as well as the presence of comorbidities (7). sRD is progressive and can lead to irreversible damage (8). While pRD is considered benign, the pain, functional disability, and unpredictability of episodes (9) lead individuals to make lifestyle changes (7). Available treatments require daily routine modifications, are often ineffective, have side effects (10), and negatively impact individuals’ quality of life and mental health (11).

Research investigating the impact of RD on quality of life is limited and has not always employed appropriate measures. Past research measured health-related quality of life using 36-Item Short Form Health Survey (SF-36; 12, 13), the anxiety/depression dimension of EQ-5D (14), and measures of wellbeing (i.e., ONS4-Life Satisfaction; 15, 16). In the absence of RD-specific quality of life measures, Fábián et al. (17) developed and validated the Raynaud Specific Quality of Life Questionnaire (RQLQ). Using RQLQ, Fábián et al. (11) reported that people with sRD had significantly lower quality of life than those with primary RD.

People with RD report higher levels of stress (15), anxiety (15, 16), and depression (11, 15, 16, 18) compared with the general population. Some studies found those with sRD being more severely affected than those with pRD (11, 16), while others found no differences between the two types (15). Variations in findings may be explained by the different measures employed by these studies. A study with people with pRD reported that stress was not a significant predictor of RD episodes; however, they found a relationship between higher anxiety and a higher frequency of episodes at temperatures over 15.5° C (19). Surprisingly, this was the only study that investigated the role of temperature on psychological outcomes.

Previous studies reported mixed outcomes between cold temperatures and RD. Maricq et al. (20) found that RD episodes were more likely to occur in colder climates. However, Pauling et al. (6) compared people with sRD in India and North America and reported that the difficulties they experienced in winter were not significantly different, despite the temperature difference between the two regions being more than 20°C. Additionally, a New Zealand study found that RD was more prevalent in the warmer region of the country (5). The relationship between temperature and RD remains underexplored and warrants further investigation.

The revised Wilson and Cleary model provides a theoretical model to understand better health-related quality of life in people living with chronic disease (21). According to this model, there are four main determinants of overall quality of life including biological function (e.g., abnormal vasoconstriction), symptoms (e.g., pain), functional status (e.g., ability to hold small objects), and general health perceptions (e.g., perception of RD severity). Characteristics of the individual (e.g., age, gender) and their environment (e.g., average temperatures) influence all determinants and quality of life.

The current study was guided by the revised Wilson and Cleary model and aimed to conduct a survey-based assessment of quality of life, wellbeing, and mental health in people living with RD. The study addressed gaps in the literature by employing an RD-specific measure for quality of life, using a large international sample, and including meteorological variables in the analyses.

## Methods

### Recruitment

A cross-sectional survey was designed, following guidelines by Kelley et al. (22). Eligibility criteria included having a diagnosis or suspecting they have RD, being over 18 years old, and being fluent in English. The survey was distributed in hard copy and online via JISC between March and April 2025. Data collection continued until two consecutive days passed without any new completions. Participants were recruited through social media and relevant charities and organisations. Ethics approval was granted by the Humanities, Social, and Health Sciences Research Ethics Panel at the University of Bradford (E1301).

### Survey design

#### Demographics

Demographic questions include participants’ age, gender, ethnicity, employment, smoking habits, and location. Based on participants’ location, the following meteorological variables were recorded: average temperature and temperature variation on the day the survey was completed and the preceding four weeks, along with precipitation, humidity, dew point, wind, and air pressure for the same four-week period. The Universal Thermal Scale was used to classify participants’ locations from coldest to warmest based on the average temperatures in the preceding four weeks (23). Participants’ climate regions were classified using the Köppen–Geiger climate classification (23). The wind chill was calculated using the following formula (24):

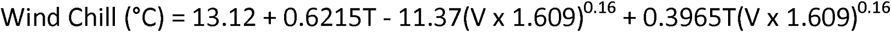

Where:

T = temperature in °C

V = wind speed in mph

#### Raynaud’s diagnosis

Participants were asked if they had been diagnosed with RD, and if yes, whether they had pRD or sRD. If participants had no diagnosis, they completed Raynaud’s quiz (25) (e.g., ‘Are your fingers or toes often cold, even indoors on a warm day and air conditioning?’) Participants provided Yes/No answers. At least one Yes suggests that the person may have RD. Total scores can range between 0 and 5.

#### Raynaud’s severity

Following previous studies (17, 19), pain severity was assessed using three questions from SF-36. These items were modified for RD. Participants were asked to report the following questions for the past 4 weeks: ‘How often did you experience pain as a result of your Raynaud’s?’ (1 = Never – 4 = Often); ‘How intense was the pain that you experienced as a result of your Raynaud’s?’ (1 = No pain – 4 = Severe pain); and ‘How much did pain as a result of your Raynaud’s interfere with your normal work (Including both outside the home and housework)?’ (1 = Not at all – 5 = Extremely). The final raw scores (ranging from 3 to 13) were rescaled to range from 5 to 100 according to the SF-36 Manual. Higher scores indicate a higher degree of pain.

Symptom severity was assessed with the question ‘How do you perceive the severity of your Raynaud’s?’. The question is rated on a 5-point Likert scale (1 = Not serious at all – 5 = Serious). Higher scores indicate higher symptom severity.

#### The Raynaud-Specific Quality of Life Questionnaire (RQLQ; 17)

RQLQ is a validated measure that assesses the impact of RD on quality of life. Participants are presented with 29 statements rated on a 5-point Likert scale (1 = Strongly agree – 5 = Strongly disagree). Scores are calculated for 5 subscales: (1) Impaired Hand Function (e.g., ‘My illness limits my grip strength’); (2) Social Interaction (e.g., ‘I have to conceal the body parts affected by my illness’); (3) Emotional Burden (e.g., ‘I am concerned that my condition will worsen)); (4) Control (e.g., ‘I wear thicker clothes); and (5) Sleep (e.g., During the night, I wake up due to numbness). Total scores can range between 29 and 145 with higher scores indicating better quality of life. Cronbach’s α for the overall scale in the current study was excellent (.94).

#### WHOIZI5 WellIZIBeing Index (WHO-5; 26)

WHO-5 is a validated measure of subjective mental wellbeing. Participants are asked five positively worded statements relating to the past two weeks (e.g., I have felt cheerful and in good spirits). Each statement is rated on a 6-point Likert scale (0 = At no time – 5 = All of the time). Total scores can range between 0 and 30 with higher scores indicating better mental wellbeing. Cronbach’s α in the current study was excellent (.90).

#### The Depression, Anxiety, and Stress Scale – 21 items (DASS-21; 27)

DASS-21 is a validated measure that assesses feelings of depression, anxiety, and stress. It comprises three 7-item scales. Participants are asked to indicate whether each statement has applied to them during the last week. Responses are given using a 4-point Likert-type scale (0lll=lllDoes not apply to me at all – 3lll=lllApplies to me a lot/most of the time). Scores are summed and multiplied by two. The total score for each scale ranges between 0 and 42. Higher scores indicate a higher level of the respective construct. The total DASS-21 score is the sum of the three scales and can range between 0 and 126 with higher scores indicating a higher level of general psychological distress. Cronbach’s α in the current study was excellent for the overall scale (.94) and Depression (.92), and good for Anxiety (.80) and Stress (.88).

### Data analysis

Statistical analyses were performed in SPSS v. 28 (28). Alpha was set at .05. Potential outliers were identified using pre-determined Z-score cut-offs (± 3.29; 29). Statistically significant outliers were identified on the RQLQ domains Emotional Burden (n = 3) and Control (n = 7), and DASS Anxiety scale (n = 4). Despite the high values, the scores were plausible and did not influence analyses.

Differences in continuous variables among the three groups were analysed using one-way ANOVA with Bonferroni correction, while Chi-square tests for independence were used to evaluate differences in categorical variables. Likelihood ratios were reported due to violations of Chi-square assumptions. Post-analysis was performed using adjusted standardised residuals.

Multivariate Analysis of Variance (MANOVA) was conducted to examine group differences in quality of life, wellbeing, stress, anxiety, and depression based on diagnosis type and climate classification. Post-hoc analyses were performed using one-way ANOVAs with Bonferroni correction. Welch’s MANOVA was reported where appropriate due to the violation of homogeneity of variance as indicated by Levene’s test. Post-hoc analyses were performed using Welch’s ANOVAs followed by Games-Howell tests. Bootstrapped MANOVAs (2,000 samples, 95% CI) were calculated to account for the non-normal distribution of scores. The assumption of homogeneity of variance-covariance matrices was assessed using Box’s M test.

Bivariate correlations assessed the relationship between quality of life, wellbeing, stress, anxiety, depression, pain and symptom severity, and meteorological variables. Spearman’s correlations were used due to violations of the assumption of normality.

Hierarchical multiple regressions were conducted separately on quality of life and wellbeing to assess the variance accounted for by type of diagnosis, pain and symptom severity, mental health, and meteorological variables. Assumptions of linearity, normality, and homoscedasticity were assessed through visual inspection of scatterplots. Models were checked for outliers (standardised residuals ± 3, Cook’s distance > 1) and multi-collinearity (tolerance < .01, variance inflation factor > 10). Bootstrapped regression analyses were run due to violations of normality.

A priori power analysis using G*Power (30) with α = .05 suggested that the sample had enough power (0.80) to detect a small effect (f = 0.02) as 628 participants were required.

## Results

A total of 721 participants completed the survey. One participant was removed (Raynaud’s quiz score was 0), leaving 720 participants in the analyses. The sample included 340 individuals with pRD, 274 individuals with sRD, and 106 individuals without a diagnosis. The three groups significantly differed on age, F(2,712) = 9.32, p < .001, η = .05, country, χ²(16) = 47.11, p < .001, Cramér’s V < .001, and employment, χ²(4) = 30.48, p < .001, Cramér’s V < .001. There were no significant differences in gender, ethnicity, and smoking (p > .05; see Table 1).

**Table 1.** Descriptive information by group.

|  | Primary RD<br>n = 340<br>n (%) | Secondary RD<br>n = 274<br>n (%) | Undiagnosed RD<br>n = 106<br>n (%) | Total sample<br>N = 720<br>N (%) | *p* |
| --- | --- | --- | --- | --- | --- |
| Age Range, *M(SD)* | 18-80, 47.22(12.97) | 20-80, 51.1(13.34) | 18-77, 45.72(13.01) | 18-80, 48.49(13.27) | < .001 |
| SF-36 *M(SD)* | 65.24(20.74) | 66.82(19.69) | 57.61(20.70) | 64.72(20.54) | < .001 |
| Gender |  |  |  |  | 1.06 |
| Female | 321 (94.4) | 258 (94.2) | 92 (86.8) | 671 (93.2) |  |
| Male | 18 (5.3) | 15 (5.5) | 12 (11.3) | 45 (6.3) |  |
| Other | 1 (.3) | 1 (.4) | 2 (1.9) | 4 (.6) |  |
| Ethnicity |  |  |  |  | .783 |
| White | 315 (92.6) | 250 (91.2) | 97 (91.5) | 662 (91.9) |  |
| Black | 2 (.6) | 2 (.7) | 1 (.9) | 5 (.7) |  |
| Asian | 8 (2.4) | 10 (3.6) | 5 (4.7) | 23 (3.2) |  |
| Mixed | 8 (2.4) | 5 (1.8) | 3 (2.8) | 16 (2.2) |  |
| Other | 4 (1.2) | 6 (2.2) | 0 (0.0) | 10 (1.4) |  |
| Country |  |  |  |  | < .001 |
| UK | 167 (49.1) | 112 (40.9) | 66 (62.3) | 345 (47.9) |  |
| USA | 131 (38.5) | 81 (29.6) | 22 (20.8) | 234 (32.5) |  |
| Canada | 15 (4.4) | 28 (10.2) | 3 (2.8) | 46 (6.4) |  |
| Australia/New Zealand | 8 (2.4) | 22 (8.0) | 3 (2.8) | 33 (4.6) |  |
| Other Europe | 10 (2.9) | 14 (5.1) | 6 (5.7) | 30 (4.2) |  |
| Other America | 3 (.9) | 1 (.4) | 3 (2.8) | 4 (.6) |  |
| Asia | 3 (.9) | 7 (2.6) | 1 (.9) | 13 (1.8) |  |
| Africa | 1 (.3) | 3 (1.5) | 2 (1.9) | 9 (1.3) |  |
| Smoking |  |  |  |  | .179 |
| Never smoked | 235 (69.1) | 165 (60.2) | 61 (57.5) | 461 (64.0) |  |
| Used to smoke | 82 (24.1) | 91 (33.2) | 36 (34.0) | 209 (29.0) |  |
| Smokes occasionally | 5 (1.5) | 5 (1.8) | 4 (3.8) | 14 (1.9) |  |
| Smokes some days | 4 (1.2) | 1 (.4) | 1 (.9) | 6 (.8) |  |
| Smokes daily | 14 (4.1) | 12 (4.4) | 4 (3.8) | 30 (4.2) |  |
| Employment |  |  |  |  | < .001 |
| Economically active | 267 (78.5) | 162 (59.1) | 82 (77.4) | 511 (71.0) |  |
| Economically inactive | 72 (21.2) | 111 (40.5) | 24 (22.6) | 207 (28.7) |  |
| Climate |  |  |  |  | .01 |
| Tropical | 4 (1.2) | 8 (2.9) | 4 (3.8) | 8 (2.9) |  |
| Dry | 21 (6.2) | 14 (5.1) | 2 (1.9) | 14 (5.1) |  |
| Temperate | 231 (67.9) | 183 (66.8) | 83 (78.3) | 183 (66.8) |  |
| Continental | 78 (22.9) | 63 (23.0) | 15 (14.2) | 63 (97.8) |  |
| Symptom severity |  |  |  |  | < .001 |
| Serious | 28 (7.8) | 20 (7.2) | 2 (1.7)* | 50 (6.6) |  |
| Quite serious | 54 (15.1) | 67 (24.0)** | 12 (10.1) | 133 (17.6) |  |
| Moderately serious | 120 (33.5) | 111 (39.8) | 31 (26.1) | 262 (34.7) |  |
| Mildly serious | 102 (33.5) | 69 (24.7)* | 36 (30.3) | 207 (27.4) |  |
| Not serious at all | 17 (5.0) | 4 (1.5) | 16 (15.1) | 37 (5.1) |  |

\* $p < .05$ , \*\* $p < .01$ . Missing responses may result in non-rounded values.

A Chi-square test revealed a significant association between type of diagnosis and symptom severity, χ²(8) = 44.11, p = .001, Cramér’s V < .001. Examination of adjusted standardized residuals indicated that participants with sRD were underrepresented in the ‘Not serious at all’ (z = – 3.70, p < .001) and ‘Mildly serious’ categories (z = – 2.10, p < .05), and overrepresented in the ‘Quite serious’ category (z = 2.9, p < .01). Participants without a diagnosis were overrepresented in the ‘Not serious at all’ category (z = 5.2, p < .001) and underrepresented in the ‘Serious category’ (z = –2.1, p < .05).

Welch’s ANOVA showed significant group differences in pain severity, F(2,290) = 7.90, p < .001, η² = .04. Games-Howell post-hoc comparison tests indicated that participants with pRD (p = .01) and sRD (p < .001) experienced a higher degree of pain compared to participants without a diagnosis.

In comparison with normative data (31), the sample had significantly higher scores on stress, t(719) = 13.41, p < .001, anxiety, t(719) = 20.74, p < .001, and depression, t(719) = 13.47, p < .001. A one-way MANOVA revealed a significant multivariate effect of type of diagnosis on quality of life, wellbeing, stress, anxiety, and depression, F(10, 1428) = 8.06, bootstrapped, p < .001, η² = .05. Follow-up Welch’s ANOVA showed significant group differences on quality of life, F(2,717) = 27.15, bootstrapped p < .001, η² = .11, and anxiety, F(2,717) = 4.18, bootstrapped p = .025, η² = .05. Univariate ANOVAs showed significant group differences on wellbeing, F(2,717) = 11.91, bootstrapped p < .001, η² = .03, stress, F(2,717) = 3.45, bootstrapped p =.03, η² = .01, and depression, F(2,717) = 6.85, bootstrapped p < .001, η² = .03 (see Table 2).

**Table 2.** Between-groups comparisons on dependent variables.

|  | Primary RD<br>n = 340<br>n (%) | Secondary RD<br>n = 274<br>n (%) | Undiagnosed RD<br>n = 106<br>n (%) | Total sample<br>N = 756<br>N (%) | *p* |
| --- | --- | --- | --- | --- | --- |
| RQLQ Total | 79.81(20.46) | 70.93(17.42) | 85.52(20.74) | 77.27(20.09) | < .001 |
| WHO-5 | 16.85(5.57) | 14.76(5.28) | 16.65(5.56) | 16.02(5.54) | <.001 |
| Stress *M(SD)* | 13.66(9.9) | 13.92(9.7) | 16.45(9.6) | 14.17(9.8) | .03 |
| Normal | 204 (60.0) | 167 (60.9) | 53 (50.0) | 424 (58.9) |  |
| Mild | 47 (13.8) | 37 (13.5) | 18 (17.0) | 102 (14.2) |  |
| Moderate | 46 (13.5) | 25 (9.1) | 16 (15.1) | 87 (12.1) |  |
| Severe | 24 (7.1) | 34 (12.4) | 8 (7.5) | 66 (9.2) |  |
| Extremely severe | 19 (5.6) | 11 (4.0) | 11 (10.4) | 41 (5.7) |  |
| Anxiety *M(SD)* | 9.45(8.2) | 10.60(9.2) | 12.13(8.5) | 10.28(8.7) | .02 |
| Normal | 157 (46.2) | 122 (44.5) | 28 (26.4) | 307 (42.6) |  |
| Mild | 47 (13.8) | 27 (9.9) | 16 (15.1) | 90 (12.5) |  |
| Moderate | 71 (20.9) | 45 (16.4) | 30 (28.3) | 146 (20.3) |  |
| Severe | 21 (6.2) | 30 (10.9) | 12 (11.3) | 63 (8.8) |  |
| Extremely severe | 44 (12.9) | 50 (18.2) | 20 (18.9) | 114 (15.8) |  |
| Depression *M(SD)* | 9.94(9.7) | 10.18(10.5) | 13.94(10.1) | 10.62(10.1) | .001 |
| Normal | 194 (57.1) | 156 (56.9) | 40 (37.7) | 390 (54.2) |  |
| Mild | 40 (11.8) | 34 (12.4) | 18 (17.0) | 92 (12.8) |  |
| Moderate | 61 (17.9) | 42 (15.3) | 25 (23.6) | 128 (17.8) |  |
| Severe | 21 (6.2) | 16 (5.8)) | 9 (8.5) | 46 (6.4) |  |
| Extremely severe | 24 (7.1) | 26 (9.5) | 14 (13.2) | 64 (8.9) |  |

The cut-off points for each scale are: for Depression, 0–9 normal, 10–13 mild, 14–20 moderate, 21–27 severe, and 28+ extremely severe; for Anxiety, 0–7 normal, 8–9 mild, 10–14 moderate, 15–19 severe, and 20+ extremely severe; and for Stress, 0–14 normal, 15–18 mild, 19–25 moderate, 26–33 severe, and 34+ extremely severe.

Post-hoc tests showed that participants with sRD had significantly lower scores than those with pRD and without a diagnosis on quality of life (both p < .001), and wellbeing (p < .001, p = .01, respectively). Participants with primary RD had significantly lower scores than those without a diagnosis on quality of life (p = .04). Participants without a diagnosis had significantly higher stress (p = 0.3) and anxiety scores (p = .01) than those with pRD, and higher depression scores than those with pRD and sRD (both p = .01).

A one-way MANOVA revealed a significant multivariate effect of climate on quality of life and wellbeing, F(15, 2100) = .046, bootstrapped p = .01 partial η² = .02. Follow-up univariate ANOVAs showed significant group differences on quality of life, F(3,702) = 4.08, bootstrapped p = .01, η² = .017, and wellbeing, F(3,702) = 3.52, bootstrapped p = .02, η² = .02. Post-hoc tests showed that participants living in tropical climates had significantly lower quality of life than those living in temperate (p = .02) and continental climates (p = .04). Additionally, participants in temperate climates had significantly lower wellbeing than those living in continental climates (p = .01).

Bivariate correlations showed that quality of life, wellbeing, pain, and symptom severity were all significantly correlated in the expected directions (Table 3). Strong correlations were found between quality of life and pain (-.551, p < .001), and symptom severity and pain (.672, p < .001). There were also strong correlations between stress and anxiety (.673, p < .001), stress and depression (.715, p < .001), and anxiety and depression (.648, p < .001; see Table 3).

**Table 3.**
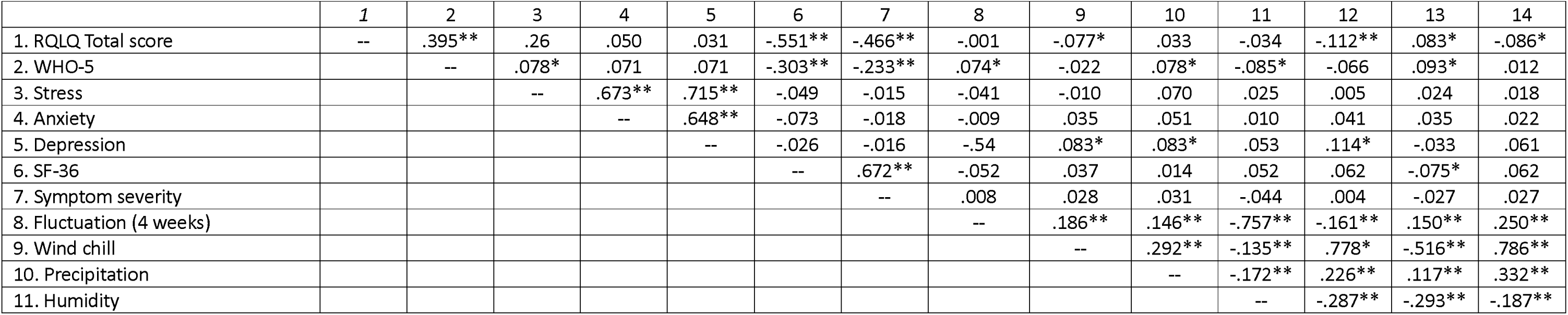
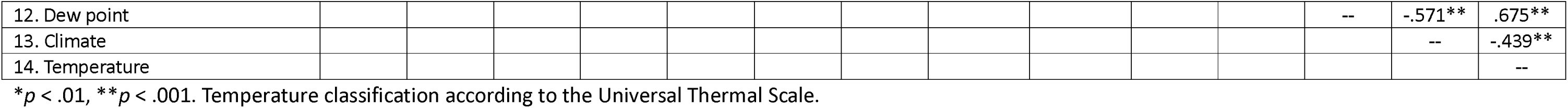
Spearman’s correlations between variables.

In Step 1, the type of diagnosis explained 7% of the variance in quality of life, R = .07, F(1,659) = 50.78, p < .001. After entry in Step 2, pain and symptom severity explained an additional 30% of the variance, R² change = .30, F change (2, 657) = 154.13, p < .001. In Step 3, wellbeing explained an additional 5% of the variance, R² change = .05, F change (1, 656) = 50.28, p < .001. In Step 4, mental health variables did not account for a significant increase in the explained variance, R² change = .001, F change (3, 656) = .20, p = .89. In Step 5, meteorological variables explained an additional 1% of the variance, R² change = .01, F change (7, 646) = 2.16, p = .04. The total variance explained by the model was 42%, R = .42, F(7,646) = 2.16, p = .04 (see Table 4).

**Table 4.**
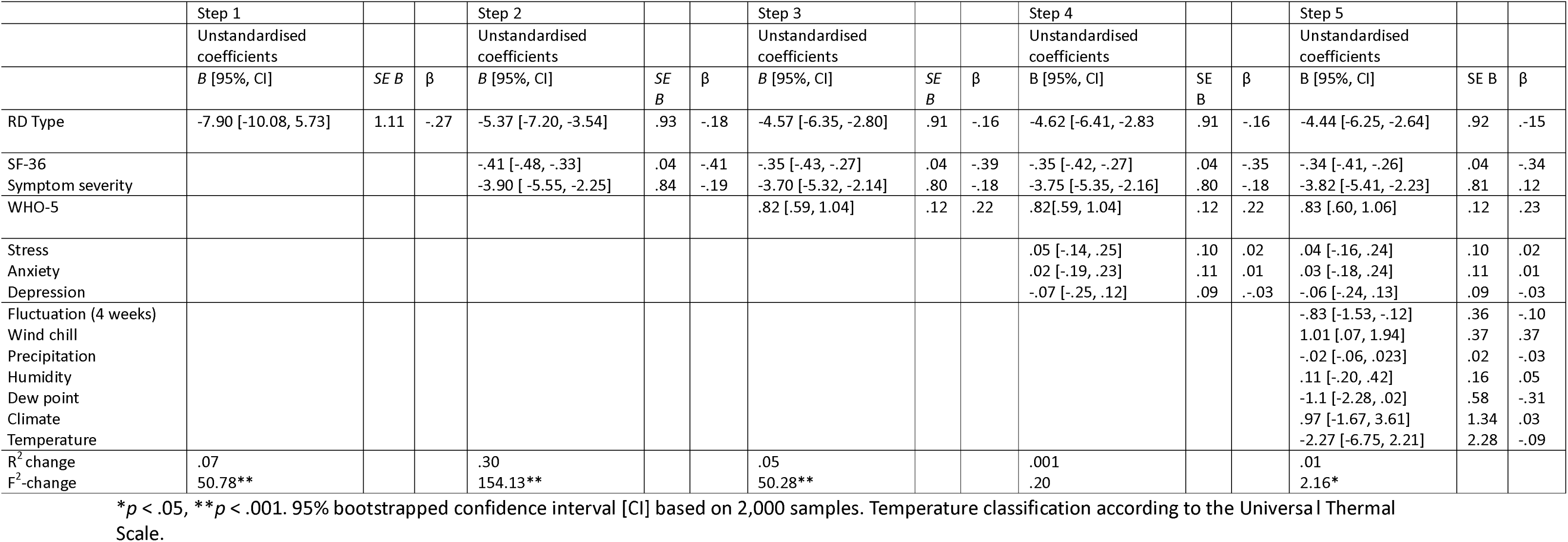
Hierarchical regression analysis for quality of life.

In Step 1, the type of diagnosis explained 3% of the variance in wellbeing, R^2^ = .03, F(1,659) = 17.39, p < .001. After entry in Step 2, pain and symptom severity explained an additional 8% of the variance, R² change = .08, F change (2, 657) = 29.01, p < .001. In Step 3, quality of life explained an additional 6% of the variance, R² change = .06, F change (1, 656) = 50.28, p < .001. In Step 4, mental health variables did not account for a significant increase in the explained variance, R² change = .004, F change (3, 656) = .10, p = .39. In Step 5, meteorological variables explained an additional 2% of the variance, R² change = .02, F change (7, 646) = 2.15, p = .04. The total variance explained by the model was 17%, R = .17, F(7,646) = 2.16, p = .04 (see Table 5).

**Table 5.**
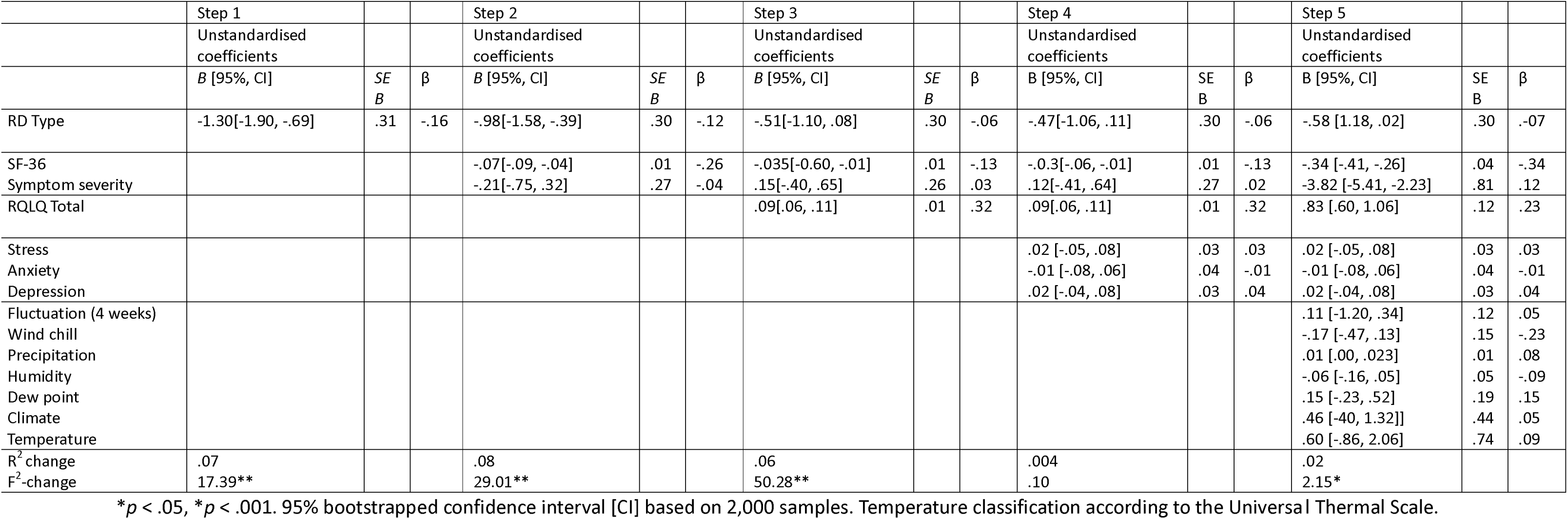
Hierarchical regression analysis for wellbeing.

## Discussion

This study assessed quality of life, wellbeing, and mental health in RD using a large, international sample. It addressed an important gap in the literature by adding various meteorological variables in the analyses and using an RD-specific measure for quality of life. It is the first study to include people without an RD diagnosis. Despite the relatively high prevalence of RD, it is frequently underdiagnosed due to a lack of diagnostic tools, lack of awareness, or symptom misinterpretation (32, 33).

The findings are consistent with a body of research showing that mental health is worse in people with RD compared with the general population (31). In line with past research participants with sRD reported lower quality of life and wellbeing (11, 15), higher anxiety and depression (11, 16), and greater pain and symptom severity than those with pRD or without a diagnosis (11, 16). A cross-sectional survey study found that people with systemic and rheumatologic diseases were amongst those reporting the poorest quality of life (34). Interestingly, participants without a diagnosis reported lower mental health than the other groups. One possible explanation is that the lack of diagnosis contributes to poor mental health. Mund et al. (35) found high rates of mental disorders in individuals seeking diagnosis for unexplained symptoms possibly due to uncertainty and associated frustration. Clinicians should prioritise early screening, diagnosis, and tailored support for people living with systemic and rheumatologic diseases.

Participants living in tropical climates reported the lowest quality of life, while those living in temperate climates reported the lowest wellbeing. These findings challenge the notion that people living in colder climates experience RD more severely. People living in warmer climates may experience more rapid temperature changes due to air conditioning. Similarly, those living in temperate climates often experience extreme weather changes or conditions, making planning and prevention difficult (36). The impact of climate on RD may be more complex than a cold/hot dichotomy. Individual (e.g., adaptive and coping strategies) and societal factors (e.g., healthcare access, infrastructure) should be considered.

Pain and symptom severity accounted for 30% of the variance in quality of life supporting the revised Wilson Cleary model, according to which symptoms and general health perceptions are determinants of overall quality of life. The strong correlation between pain and symptom severity highlights their role in influencing patients’ experiences. Mental health variables did not significantly account for an additional variance in either quality of life or wellbeing. Measures of quality of life or wellbeing may capture mental health indirectly (37) or there is an overlap between physical symptoms and mental health symptoms (38). Meteorological variables accounted for up to 2% of the variance in quality of life and wellbeing, suggesting that although they are important, physical variables are more important. Interventions targeting pain and symptoms management should be prioritised to improve the quality of life and wellbeing of people living with RD. Pain and symptom relief may also benefit mental health. While meteorological variables matter, they should complement rather than replace other assessments.

### Limitations and future research

The study’s cross-sectional design does not allow us to infer how the relationships between the variables may differ over time. Self-reported data introduce biases including subjective interpretation of symptoms. Although including participants without a diagnosis is important given the underdiagnosis of RD, it is possible that some of those included did not have RD. Recruitment through social media and charities may have attracted those who are more symptomatic or more engaged in RD campaigns reducing the generalisability of findings.

Several other variables may have affected the outcomes and should be included in future studies (e.g., healthcare access, coping strategies). Future studies should also consider longitudinal designs to establish the directionality of influences and include clinical samples.

### Conclusion

This study showed that diagnosis type, pain, and symptom severity significantly affect quality of life, wellbeing, and mental health, while meteorological variables play a smaller role. Individuals with sRD experience the greatest burden including greater pain, and poorer quality of life and mental health. Tailored interventions targeting pain and symptom severity, are critical for improving patients’ quality of life and wellbeing. Climate influences should be considered alongside clinical and psychosocial factors in healthcare planning.

## Data Availability Statement

The data underlying this article will be shared on reasonable request to the corresponding author.

